# Association of preoperative plasma suPAR levels with intraoperative sublingual microvascular perfusion in patients undergoing major non-cardiac surgery

**DOI:** 10.1101/2021.07.03.21259960

**Authors:** Athanasios Chalkias, Nikolaos Papagiannakis, Bernd Saugel, Moritz Flick, Konstantina Kolonia, Zacharoula Angelopoulou, Dimitrios Ragias, Dimitra Papaspyrou, Nicoletta Ntalarizou, Aikaterini Bouzia, Konstantinos Stamoulis, Aikaterini Kyriakaki, Jesper Eugen-Olsen, Eleni Laou, Eleni Arnaoutoglou

## Abstract

**INTRODUCTION:** The plasma suPAR level has previously been associated with postoperative complications and has been shown to be an independent predictor of coronary microvascular function and flow reserve. We investigated the association between preoperative suPAR levels and intraoperative sublingual microvascular perfusion in patients undergoing elective major non-cardiac surgery.

**Methods:** This study included 100 patients undergoing major non-cardiac surgery between February 2019 and September 2020. The primary objective was to investigate the association between preoperative suPAR and intraoperative sublingual De Backer score, Consensus Proportion of Perfused Vessels (Consensus PPV), and Consensus PPV (small). Secondary objectives were to investigate the associations between these sublingual microcirculatory variables and (1) complications and (2) mean arterial pressure. EDTA blood was collected before induction of anesthesia and plasma suPAR levels were determined using the suPARnostic® quick triage lateral flow assay. Sublingual microcirculation was monitored with Sidestream DarkField (SDF+) imaging technique at 20 minutes after induction of anesthesia before surgical incision (baseline) and then every 30 minutes until emergence from anesthesia.

**Results:** A decrease of 0.7 mm^-1^ in the De Backer score, 2.5% in the Consensus PPV, and 2.8% in the Consensus PPV (small) from baseline measurement was observed for every 1 ng/ml increase of suPAR or 1 additional minute of intraoperative time. De Baker score did not change significantly from baseline (p=0.404), while Consensus PPV and Consensus PPV (small) decreased significantly from baseline (p<0.001 in both cases). The De Backer score, the Consensus PPV, and the Consensus PPV (small) correlated with postoperative complications. Mean arterial pressure correlated with De Backer score (p=0.487) but not with Consensus PPV (p=0.506) or Consensus PPV (small) (p=0.697) during the intraoperative period.

**Conclusion:** Preoperative suPAR levels and prolonged operative duration were associated with intraoperative impairment of sublingual microvascular perfusion in patients undergoing elective major non-cardiac surgery.

## INTRODUCTION

The microcirculation is the terminal vascular network of the systemic circulation and is key for oxygen transfer to the parenchymal cells. Our understanding of the microcirculation has evolved over the last decades and it has been shown that the microcirculation can be altered even when systemic hemodynamics are considered normal.^1,2^ Hand-held vital microscopes can be used to observe sublingual microcirculatory alterations at the bedside. The sublingual microcirculation reflects splanchnic microcirculatory perfusion and thus provides physiological insight helping to achieve a more physiological approach to perioperative management.^3,4^

There is evidence suggesting an association between chronic systemic inflammation and impaired microcirculatory perfusion. A prolonged inflammatory response may impair vasomotor function and/or increase the rate of proliferation of blood vessels, eventually reducing capillary perfusion and leading to organ dysfunction.^5,6^ In turn, the inflammation-induced microvascular changes may enhance and sustain the inflammatory response.^1,5^

The soluble form of urokinase plasminogen activator receptor (suPAR) is a marker for systemic inflammation. The specific physiologic role of suPAR is unclear, but its levels in circulation reflect the chronic activation of the inflammatory and immune systems.^7,8^ Compared to C-reactive protein, suPAR is a more stable molecule, which makes it a more reliable biomarker for reflecting the state of chronic immune activation of the patient.^9^ Not only suPAR is implicated in the inflammatory response of several diseases, but it was recently reported that individuals can be also genetically predisposed to higher or lower suPAR levels, which give suPAR a significant prognostic value in physiological or pathological conditions.^10^

We have recently shown that the preoperative suPAR levels are highly predictive of postoperative complications in different types of surgical procedures or different risk groups,^11^ but the reasons for this are unknown. Interestingly, the suPAR level has previously been demonstrated to be an independent predictor of coronary microvascular dysfunction in patients with non-obstructive coronary artery disease, with doubling of plasma suPAR being associated with about 30% decrease in coronary flow reserve and, eventually, with heart disease and worse long-term outcomes.^12,13^ We hypothesized that chronic systemic inflammation, reflected by preoperative suPAR levels, is associated with impaired intraoperative microvascular perfusion in patients having non-cardiac surgery. To test this hypothesis, we performed a prospective observational study and investigated the association between preoperative suPAR levels and intraoperative sublingual microvascular perfusion in patients undergoing elective major non-cardiac surgery.

## METHODS

### Design

This analysis included 100 patients of a previous prospective observational study conducted in the University Hospital of Larisa, Larisa, Greece, from February 2019 to September 2020.^11^ Ethical approval for this study was provided by the Ethical Committee of the university hospital of Larisa, Greece (IRB no. 60580). The study was designed in accordance with the declaration of Helsinki and was registered at ClinicalTrials.gov (NCT03851965). Written informed consent was obtained from all patients.

### Study objectives

The primary objective was to investigate the association between preoperative suPAR and intraoperative sublingual De Backer score, Consensus Proportion of Perfused Vessels (Consensus PPV), and Consensus PPV (small). The De Backer score is based on the principle that density of the vessels is proportional to the number of vessels crossing arbitrary lines.^14,15^ It is calculated as the number of vessels crossing the lines divided by the total length of the lines. In addition, the vessel crossings are checked for flow to calculate Perfused De Backer density. Consensus PPV is then calculated as a percentage ratio of the Perfused De Backer density and De Backer density. These calculations are carried out for all vessels and small (up to 20 microns in diameter) vessels (Consensus PPV (small)). Secondary objectives were to investigate the associations between these sublingual microcirculatory variables and (1) complications and (2) mean arterial pressure (MAP).

### Patient eligibility

Consecutive patients who were scheduled to undergo elective major non-cardiac surgery with an expected duration of ≥2 hours under general anesthesia were screened for inclusion. All operative approaches were eligible for inclusion, including open and laparoscopic procedures. Patients fulfilling the following criteria were included: age ≥18 years and American Society of Anesthesiologists’ (ASA) physical status I to IV.

We excluded patients with any infection within the previous four weeks; severe liver disease; need for renal replacement therapy; allergies; inflammatory or immune system disorders; asthma; obesity (BMI ≥30 kg m^-2^); mental disability or severe psychiatric disease; alcohol abuse; connective tissue disease including rheumatoid arthritis, ankylosing spondylitis, and systemic lupus erythematosus. We also excluded patients who had previously received an organ transplant; who were treated with steroids, antipsychotic or anti-inflammatory/immunomodulatory medication within the previous three months or with opioids during the past week; and who were involved in another study. Legal incapacity or limited legal capacity were also exclusion criteria.

### Anesthetic management

Induction of anesthesia was in supine position and included i.v. midazolam 0.15-0.35 mg/kg over 20-30 seconds, fentanyl 1 μg/kg, ketamine 0.2 mg/kg, propofol 1.5-2 mg/kg, rocuronium 0.6 mg/kg, and a fraction of inspired oxygen of 70%. All drugs were prepared in labelled syringes and induction was achieved by administration of a predetermined i.v. bolus dose considering the patient’s weight and/or age. Laryngoscopy and intubation proceeded in a standard fashion, while the position of the endotracheal tube was confirmed by auscultation and capnography/capnometry. The patients were then connected to an automated ventilator (Draeger Perseus A500®; Drägerwerk AG & Co., Lübeck, Germany) and were ventilated using a lung-protective strategy with tidal volume of 7 mL/kg, positive end-expiratory pressure of 6-8 cmH_2_O, and plateau pressures <30 cmH_2_O.

Maintenance of general anesthesia initially included desflurane 1.0 MAC with 40% oxygen and 60% air. Desflurane was used for maintenance because it produces stable effects on the microcirculation compared to sevoflurane.^16^ Depth of anesthesia (bispectral index-BIS, Covidien, France) was monitored, with the target ranging between 40 and 60.^17,18^ Intraoperative fraction of inspired oxygen was then adjusted to maintain an arterial oxygen partial pressure of 80-100 mmHg^19-21^ and normocapnia was maintained by adjusting the respiratory rate as needed. Normothermia (37 °C) and normoglycemia were maintained during the perioperative period. Vasoactive drugs were administrated, if necessary, to maintain a MAP of 65 mmHg but were not provided 10 minutes prior to or during the assessment of microcirculation. After induction of anesthesia, the patients were allowed to stabilize for 20 minutes.^19,21^

### Sampling and laboratory measurements

Participants underwent sampling of peripheral venous blood immediately after arrival to the operating room before induction of anesthesia. Blood samples drawn from all patients were collected in EDTA tubes and were centrifuged at 3.000 x g for 1 minute. According to the manufacturer’s instructions, there is no detectable impact on plasma suPAR concentration when comparing 1 and 10 minutes of centrifugation. Plasma suPAR levels were then determined using the suPARnostic® Quick Triage lateral flow assay (ViroGates, Denmark). This is an easy-to-use quantitative test that is based on the lateral flow principle. The device consists of a nitrocellulose membrane with two immobilized antibody zones and a running buffer with gold particles. The quantitative results are read within 20 minutes by an optical aLF Reader (Qiagen, Germany).^11^ suPAR levels higher than 5.5 ng/ml were considered as a clinically important indicator of systemic inflammation. We chose the cut-off of 5.5 ng/ml as this has been previously used in a study of preoperative suPAR levels and post-operative complications and thus allows for comparison of previous findings.^22^ As the chosen cut-off gave a rather large group above 5.5 ng/ml, we chose a second cut-off at 10 ng/ml.^11^ There is no specific rationale for this second cut-off, except that suPAR in double digits is often referred to unusually high levels.

### Sublingual microcirculation analysis

Sublingual microcirculation was monitored using the sidestream dark field (SDF+) imaging technique (Microscan; Microvision Medical BV, Amsterdam, The Netherland). Microscan is equipped with a ring of green light-emitting diodes located at the end of a probe using a wavelength of 540 nm that is absorbed by the hemoglobin contained in red blood cells.^15,23^

Videos from at least five different sites of sublingual microcirculation were recorded. The baseline measurement was performed 20 minutes after induction of general anesthesia before surgical incision and then every 30 minutes until emergence from anesthesia. All videos were recorded by the same investigator and significant efforts were made to avoid pressure and movement artefacts, improve focus and illumination, and clean the sublingual mucosa from saliva and/or blood. Before analysis, all videos were evaluated by two experienced raters according to a modified micro-circulation image quality score (MIQS).^24^ The best three videos from each recording were analysed offline by a blinded investigator with the AVA4.3C Research Software (Microvision Medical, Amsterdam, NL).^25,26^ We used the De Backer score as density score and the Consensus PPV and the Consensus PPV (small) as flow scores.^27^

### Assessment of postoperative complications

We used the Clavien-Dindo Classification to assess postoperative complications, morbidity, and mortality in our patients.^28,29^ The Comprehensive Complication Index (CCI) calculator is an online tool to support the assessment of patients’ overall morbidity (https://www.assessurgery.com/clavien-dindo-classification/). The CCI is based on the complication grading by Clavien-Dindo Classification and the overall morbidity is reflected on a scale from 0 (no complication) to 100 (death).

This scoring system offers the advantages of being able to compare results over different time periods within the same institution.^28^ According to related studies, a CCI of 26.2 was set as the cut-off point (equivalent to one grade IIIa complication by the Clavien-Dindo Classification) and patients with complications were divided into a high-CCI group (group A, CCI ≥ 26.2) and a low-CCI group (group B, CCI < 26.2) accordingly.^30^

### Data collection, monitoring, and management

Data analysis was based on predefined data points on a prospective data collection form. The staff was blinded to measurements until the end of the study and all data were analyzed. Clinical monitoring throughout the study was performed to maximize protocol adherence, while an independent Data and Safety Monitoring research staff monitored safety, ethical, and scientific aspects of the study. Data collection included demographics, ASA score, anesthesia variables, general blood count, biochemistry profile, suPAR, and C-reactive protein. Considering that the ASA score is not designed to predict mortality, has known inter-rater variation, and offers at least a moderate predictive ability for mortality in multiple surgical settings, we also included the Modified Frailty Index, POSSUM, and ACS-NSQIP risk scores in our analysis for the purposes of perioperative risk adjustment.^31^

The goal of the clinical data management plan was to provide high-quality data by adopting standardized procedures to minimize the number of errors and missing data, and consequently, to generate an accurate database for analysis. Remote monitoring was performed to signal early aberrant patterns, issues with consistency, credibility, and other anomalies. Any missing and outlier data values were individually revised and completed or corrected whenever possible.

### Statistical analysis

Statistical analysis was performed using R v4.0. Spearman’s method was used to correlate microcirculation indices with participants’ hemodynamic and clinical characteristics, and morbidity and mortality scores. The Benjamini-Hochberg false discovery rate correction was applied in the resulting P values to account for the multiple numbers of tests. Adjusted p-values less than 0.05 were deemed significant. Linear mixed effects models with Restricted Maximum Likelihood Estimation (REML) were used in order to assess the different effects in microcirculatory indices. LME was used instead of repeated-measures ANOVA, due to the different intra-operative duration between patients and the associated missing values. Three linear mixed models were constructed, for each one of the microcirculatory variables (De Backer score, Consensus PPV, and Consensus PPV (small)), with suPAR and the different time points as fixed factors, and the different subjects (patients) as random factors. Changes in intraoperative sublingual microcirculatory variables were assessed by the second term of the model. We chose to include 100 individuals as we expected that this number could reveal important associations and generate results to be used for sample size estimation in future large-scale studies.

## RESULTS

Overall, 100 patients were included in the study, of whom 68 (68%) were men and 32 (32%) were women, with a median age of 70 years (IQR 62.5-75.5). Demographic and clinical characteristics are shown in Table 1 and Supplementary Table 1.

**Table 1.**
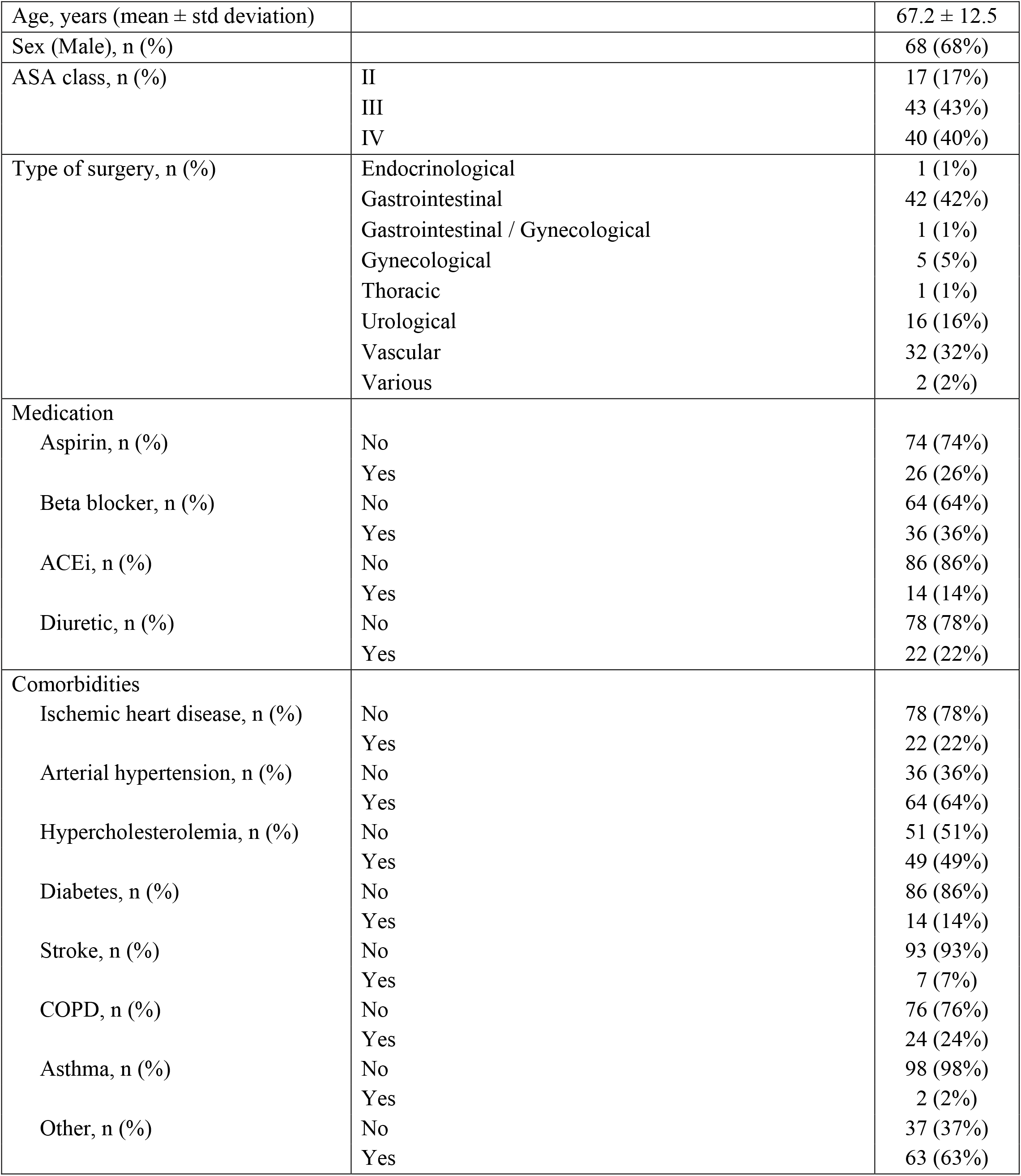
Demographic and clinical characteristics of patients undergoing major non-cardiac surgery.

|  |  |  |
| --- | --- | --- |
| Age, years (mean ± std deviation) | | 67.2 ± 12.5 |
| Sex (Male), n (%) |  | 68 (68%) |
| ASA class, n (%) | II | 17 (17%) |
|  | III | 43 (43%) |
|  | IV | 40 (40%) |
| Type of surgery, n (%) | Endocrinological | 1 (1%) |
|  | Gastrointestinal | 42 (42%) |
|  | Gastrointestinal / Gynecological | 1 (1%) |
|  | Gynecological | 5 (5%) |
|  | Thoracic | 1 (1%) |
|  | Urological | 16 (16%) |
|  | Vascular | 32 (32%) |
|  | Various | 2 (2%) |
| Medication |  |  |
| Aspirin, n (%) | No | 74 (74%) |
|  | Yes | 26 (26%) |
| Beta blocker, n (%) | No | 64 (64%) |
|  | Yes | 36 (36%) |
| ACEi, n (%) | No | 86 (86%) |
|  | Yes | 14 (14%) |
| Diuretic, n (%) | No | 78 (78%) |
|  | Yes | 22 (22%) |
| Comorbidities |  |  |
| Ischemic heart disease, n (%) | No | 78 (78%) |
|  | Yes | 22 (22%) |
| Arterial hypertension, n (%) | No | 36 (36%) |
|  | Yes | 64 (64%) |
| Hypercholesterolemia, n (%) | No | 51 (51%) |
|  | Yes | 49 (49%) |
| Diabetes, n (%) | No | 86 (86%) |
|  | Yes | 14 (14%) |
| Stroke, n (%) | No | 93 (93%) |
|  | Yes | 7 (7%) |
| COPD, n (%) | No | 76 (76%) |
|  | Yes | 24 (24%) |
| Asthma, n (%) | No | 98 (98%) |
|  | Yes | 2 (2%) |
| Other, n (%) | No | 37 (37%) |
|  | Yes | 63 (63%) |

### Association of suPAR with intraoperative sublingual microcirculatory variables

Elevated levels of preoperative suPAR were associated with a decrease in all intraoperative sublingual microcirculatory variables at all time points. Specifically, we observed a decrease of 0.7 mm^-1^ in the De Backer score, 2.5% in the Consensus PPV, and 2.8% in the Consensus PPV (small) from baseline measurement for every 1 ng/ml increase of suPAR or 1 additional minute of intraoperative time (Table 2).

**Table 2.** Association of suPAR with intraoperative sublingual microcirculatory perfusion.

|  | **Beta coefficient <sup>a</sup>** | **Standard Error** | **p-value** |
| --- | --- | --- | --- |
| **De Baker score** |  |  |  |
| suPAR | -0.716 | 0.041 | <0.001 |
| Time (min) | 0.0004 | 0.0004 | 0.405 |
| **Consensus PPV** |  |  |  |
| suPAR | -2.490 | 0.159 | <0.001 |
| Time (min) | -0.011 | 0.002 | <0.001 |
| **Consensus PPV (small)** |  |  |  |
| suPAR | -2.835 | 0.1713 | <0.001 |
| Time (min) | -0.0102 | 0.0019 | <0.001 |

<sup>a</sup> Represents the increase/decrease in the respective microcirculatory variable for every 1 ng/ml increase of suPAR or 1 additional minute of intraoperative time.

Table 3 and Figure 1 depict the estimated differences between different time points and baseline (30 min). The overall test for differences in microcirculatory variables revealed that De Baker score did not change significantly from baseline (p=0.404), while Consensus PPV and Consensus PPV (small) decreased significantly from baseline (p<0.001 in both cases) during the intraoperative period (Supplementary Table 2).

**Table 3.**
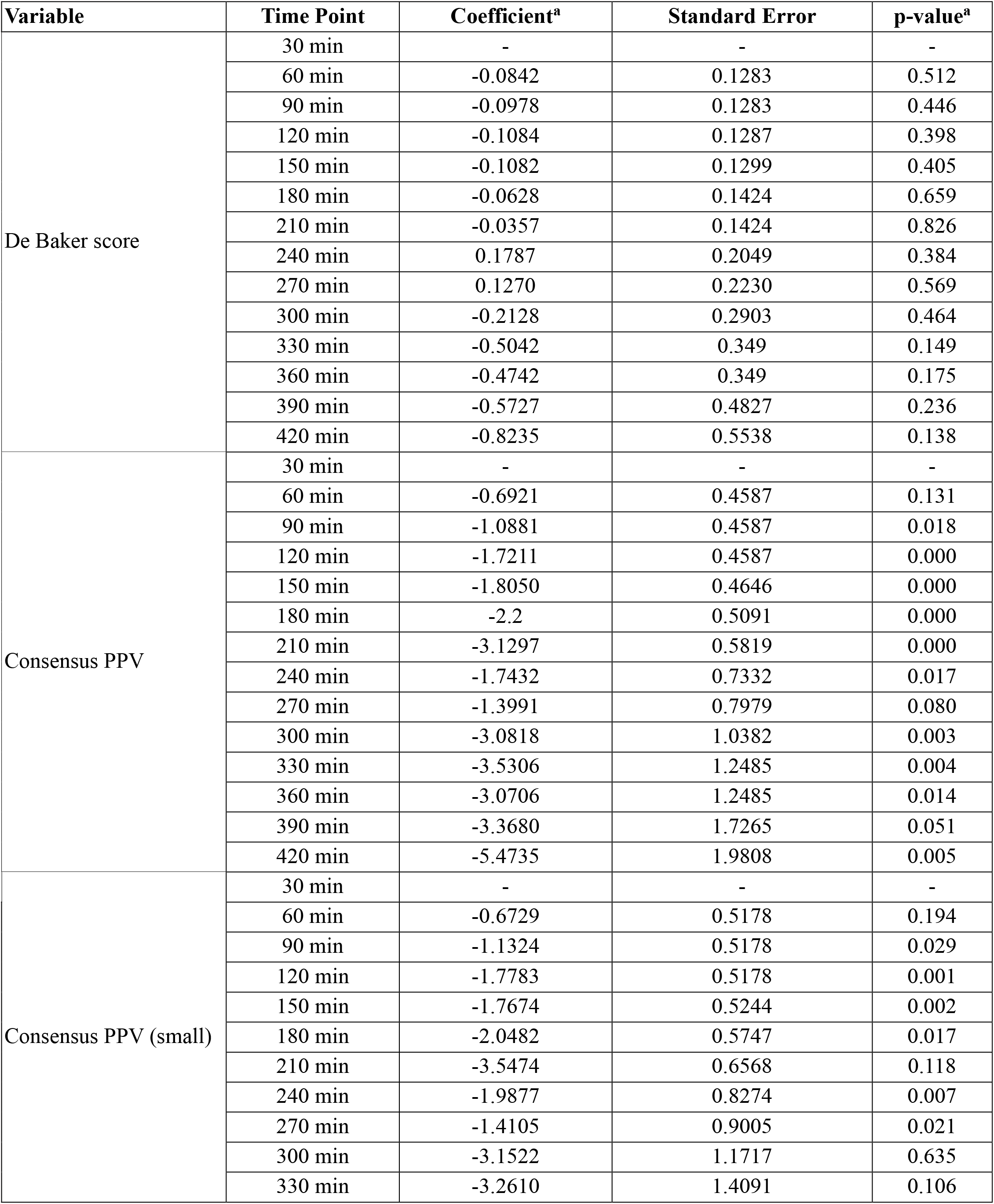
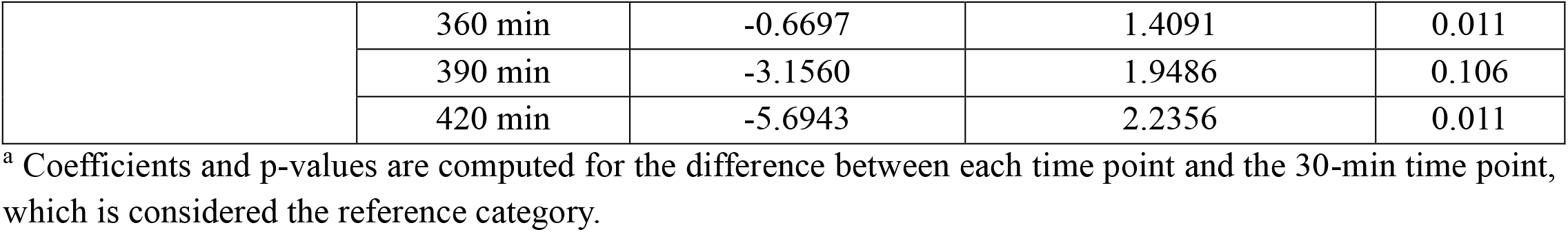
Estimated differences between different intraoperative time points and baseline (30 min)

**Figure 1.**
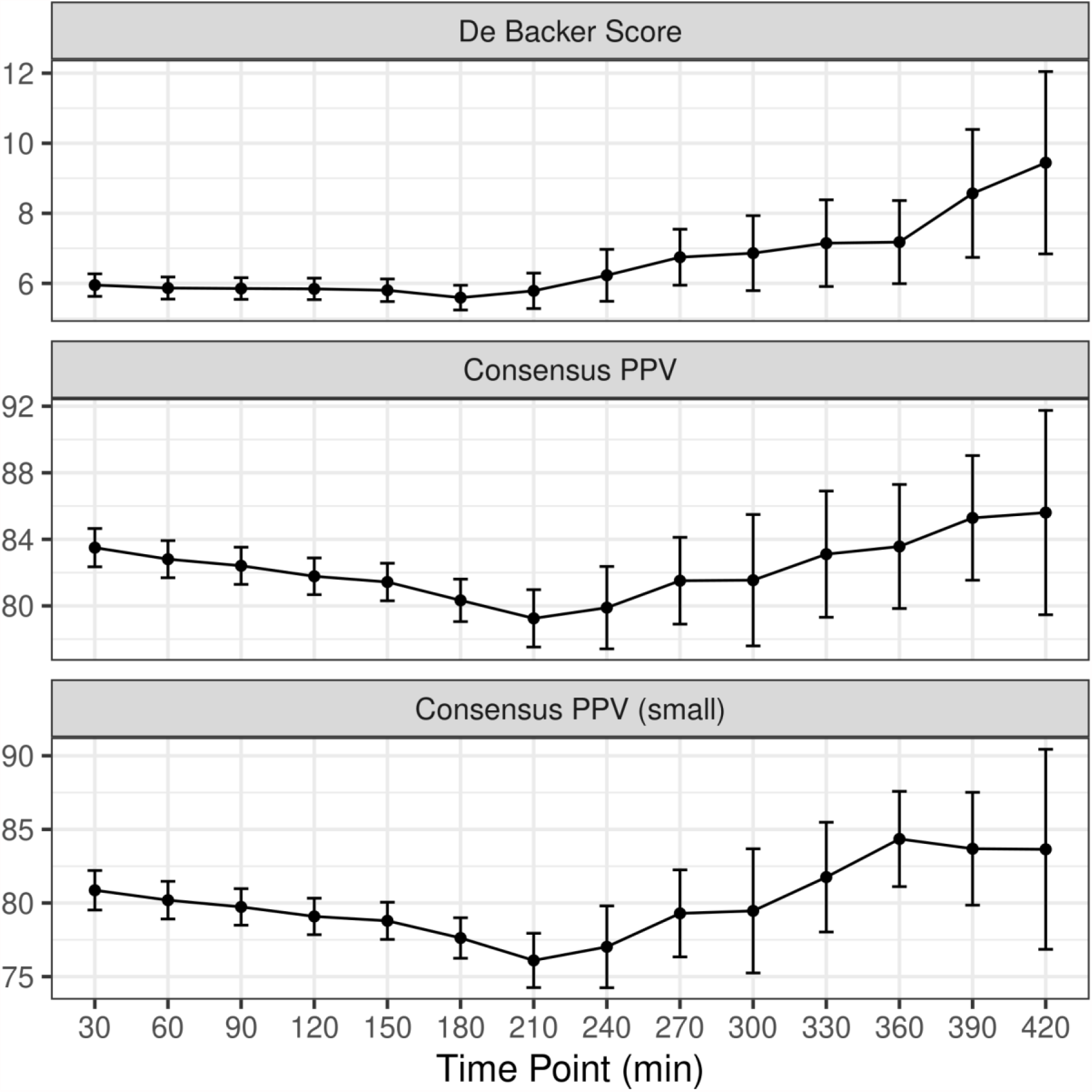
Change in De Backer score (mm^-1^), Consensus PPV (%), and Consensus PPV (small) (%) with time.

### Secondary outcomes

The De Backer score, the Consensus PPV, and the Consensus PPV (small) correlated with CCI and POSSUM risk score (Table 4). Mean arterial pressure correlated with De Backer score during the intraoperative period, but the correlation was weak (p=0.487). Mean arterial pressure did not correlate with Consensus PPV (p=0.506) or Consensus PPV (small) (p=0.697) at any time point during the intraoperative period (Supplementary Figures 1-3, Supplementary Table 3).

**Table 4.** Correlation of microcirculation variables with morbidity and mortality scores.

| **Preoperatively** |  |  |  |
| --- | --- | --- | --- |
|  |  | **Spearman's rho** | **Adjusted p-value** |
| De Backer score | Modified Frailty Index | -0.156 | 0.12 |
|  | POSSUM morbidity score | -0.224 | **0.025** |
|  | POSSUM mortality score | -0.226 | **0.024** |
|  | ACS-NSQIP score | -0.144 | 0.153 |
|  | ACS-NSQIP excess score | -0.000267 | 0.998 |
|  | Comprehensive Complication Index | -0.251 | **0.012** |
| Consensus PPV (%) | Modified Frailty Index | -0.102 | 0.315 |
|  | POSSUM morbidity score | -0.26 | **0.009** |
|  | POSSUM mortality score | -0.257 | **0.01** |
|  | ACS-NSQIP score | -0.149 | 0.138 |
|  | ACS-NSQIP excess score | 0.024 | 0.816 |
|  | Comprehensive Complication Index | -0.28 | **0.005** |
| Consensus PPV (small) (%) | Modified Frailty Index | -0.075 | 0.456 |
|  | POSSUM morbidity score | -0.223 | **0.025** |
|  | POSSUM mortality score | -0.222 | **0.027** |
|  | ACS-NSQIP score | -0.179 | **0.074** |
|  | ACS-NSQIP excess score | 0.031 | 0.759 |
|  | Comprehensive Complication Index | -0.277 | **0.005** |
| **Postoperatively** |  |  |  |
|  |  | **Spearman's rho** | **Adjusted p-value** |
| De Backer score | Modified Frailty Index | -0.184 | 0.067 |
|  | POSSUM morbidity score | -0.262 | **0.009** |
|  | POSSUM mortality score | -0.265 | **0.008** |
|  | ACS-NSQIP score | -0.141 | 0.162 |
|  | ACS-NSQIP excess score | -0.121 | 0.232 |
|  | Comprehensive Complication Index | -0.289 | **0.003** |
| Consensus PPV (%) | Modified Frailty Index | -0.221 | **0.027** |
|  | POSSUM morbidity score | -0.324 | **0.001** |
|  | POSSUM mortality score | -0.322 | **0.001** |
|  | ACS-NSQIP score | -0.128 | 0.206 |
|  | ACS-NSQIP excess score | -0.175 | 0.082 |
|  | Comprehensive Complication Index | -0.328 | **0.001** |
| Consensus PPV (small) (%) | Modified Frailty Index | -0.189 | 0.06 |
|  | POSSUM morbidity score | -0.29 | **0.003** |
|  | POSSUM mortality score | -0.289 | **0.003** |
|  | ACS-NSQIP score | -0.14 | 0.165 |
|  | ACS-NSQIP excess score | -0.164 | 0.104 |
|  | Comprehensive Complication Index | -0.327 | **0.001** |

## DISCUSSION

In this observational study with patients undergoing elective major non-cardiac surgery, we observed a decrease of 0.7 mm^-1^ in the De Backer score, 2.5% in the Consensus PPV, and 2.8% in the Consensus PPV (small) from baseline measurement for every 1 ng/ml increase of suPAR or 1 additional minute of intraoperative time. The De Baker score did not change significantly from baseline, while the Consensus PPV and the Consensus PPV (small) decreased significantly from baseline during the intraoperative period. Our findings indicate an association between the level of preoperative systemic inflammation and intraoperative impairment of sublingual microvascular perfusion, which may be particularly important during prolonged operation/anesthetic times.

Chronic inflammation is a major problem in the general population due to environmental and lifestyle factors.^32^ The level of chronic inflammation is typically assessed using biomarkers of acute inflammation, such as C-reactive protein, white blood cell counts, or proinflammatory cytokines. However, these are rapidly up-and down-regulated, as their biological function is to tightly control the acute inflammatory response, making their quantification time-sensitive. In recent years, suPAR has been investigated as a more stable biomarker of chronic inflammation, and elevated suPAR is closely linked to accelerated biological ageing, cognitive impairment, and development of inflammation-related diseases.^33-35^ Of note, the higher suPAR level decreases the function of the urokinase plasminogen activator receptor system in endothelial cells and may lead to microcirculatory flow abnormalities.^36^ In the present study, we observed a decrease in the De Backer score, the Consensus PPV, and the Consensus PPV (small) for every 1 ng/ml increase of preoperative suPAR. To the best of our knowledge, this is the first report of the inverse correlation between chronic systemic inflammation, reflected by increased preoperative suPAR levels, and sublingual microcirculatory variables in perioperative medicine. Interestingly, evidence from other populations show that increasing suPAR levels are associated with a more severe microvascular involvement,^12,36^ which is agreement with our findings.

The induction of an inflammatory response can impair the ability of microvasculature network to coordinate a vasodilatory response in different organs, especially in hypertensive individuals.^37^ suPAR is a selective inhibitor of plasminogen activator induced dilation and inhibits urokinase plasminogen activator-mediated vasodilation without compromising its catalytic activity.^38,39^ Also, inflammation and hypoxia/ischemia upregulate urokinase plasminogen activator, which impairs hypercapnic and hypotensive dilation.^40,41^ A diminished microcirculatory blood flow may in turn enhance the inflammatory response,^1,5,42^ forming a dangerous vicious cycle. The crucial point for this seems to be the change of the endothelium from a quiescent into an active state. In patients with chronic inflammation, the endothelium has been activated much earlier than the time of surgery and is associated with elevated suPAR levels.^12,43^ In these individuals, graded changes in venular shear rate even for brief periods elicit progressive recruitment of both rolling and firmly adherent leukocytes.^44,45^ The activated leukocytes cause oxidative and enzymatic degradation of the glycocalyx and further impair microvascular perfusion.^46,47^ All these are consistent with our findings, which indicate a strong association between suPAR and microcirculatory flow impairment.

The evidence suggesting an association between microvascular disease and outcomes after non-cardiac surgery is scarce. In a prospective observational study evaluating the relationship between global oxygen delivery, microcirculatory flow, and tissue oxygenation, impairment of microcirculatory flow was more marked in those patients who developed complications.^48^ Of note, the likelihood of developing a complication increases with increasing operative time, but the exact mechanisms underlying this positive association are not fully understood.^49-51^ We herein report that the De Backer score, the Consensus PPV, and the Consensus PPV (small) were inversely correlated with the CCI and the POSSUM risk score, which is important considering that a further decrease in all microcirculatory variables was observed for every 1 ng/ml increase of suPAR or 1 additional minute of intraoperative time. The CCI adds information on postoperative morbidity, with particular value following extensive surgery and longer postoperative observation periods,^29^ while the POSSUM is calculated preoperatively and has been modified and validated for numerous subtypes of surgeries and clinical scenarios. Given that the sublingual microcirculatory alterations occurring during non-cardiac surgery may be maintained during the immediate and early postoperative period,^46,48,52^ the cumulative effects of chronic inflammation and prolonged operative time on microvascular perfusion are important in the occurrence of postoperative complications.

The impairment of microcirculatory perfusion may be independent of systemic hemodynamics.^43,52,53^ This was also evident in this study, which revealed only a weak correlation between MAP and De Backer score during the intraoperative period. De Backer et al. reported that the microcirculatory perfusion might only deteriorate when MAP is lower than 60-65 mmHg.^54,55^ Maintaining MAP at higher targets may be more effective for improving splanchnic perfusion during surgery, but this may require the use of exogenous vasopressors which increase systemic vascular resistance and may aggravate microcirculatory perfusion.^54-56^ In the present study, intraoperative MAP was maintained 60-120 mmHg and therefore, we can argue that the inflammation-induced uncoupling of systemic and sublingual microcirculatory variables may not be corrected within this MAP range. Further research is necessary to clarify the effects of MAP on sublingual microcirculatory perfusion.

The inclusion of many patients with ASA classifications III and IV allowed to investigate changes of sublingual microcirculatory variables in high-risk patients and their correlation with suPAR. The main limitation is that it is a single-center study and should be reproduced in a multicenter study to improve general applicability. In this study, microcirculatory variables were determined in the sublingual vascular bed. We used the sublingual space as the site of imaging because it shares the same embryologic origin as the splanchnic mucosa and can reflect derangements in splanchnic blood flow.^3,4^ However, regional sublingual microcirculation may not always reflect regional microcirculation in vital organs. Also, the software used for quantification of sublingual microcirculatory perfusion is not validated at this time. The number of patients may be relatively small for the different types of surgical interventions, while the absence of significant differences for some study variables may be due to the sample size. However, this is one of the largest prospective cohorts investigating the correlation between chronic systemic inflammation and sublingual microcirculatory perfusion in perioperative medicine. Another limitation is that 36% and 14% of the patients received β-blockers and angiotensin-converting enzyme inhibitors, respectively. Nevertheless, both were stopped 24 hours before surgery and thus, their impact is minimal.^57^ Despite the limitations, the present study revealed important associations and results that can be used in future studies.

## CONCLUSION

Preoperative suPAR levels and prolonged operative duration were associated with intraoperative impairment of sublingual microvascular perfusion in patients undergoing elective major non-cardiac surgery. These findings may help identifying patients that may benefit from advanced perioperative care planning.

## Data Availability

Data from SPARSE can be made available upon request through a collaborative process. Please contact the corresponding author for additional information.

## Conflicts of interest

Dr. Jesper Eugen-Olsen is a co-founder, shareholder and CSO of ViroGates A/S and is mentioned inventor on patents on suPAR owned by Copenhagen University Hospital Hvidovre, Denmark. Dr. Bernd Saugel is a consultant for and has received honoraria for giving lectures from Edwards Lifesciences Inc. (Irvine, CA, USA) outside the submitted work. Dr. Bernd Saugel is a consultant for and has received institutional restricted research grants and honoraria for giving lectures from Pulsion Medical Systems SE (Feldkirchen, Germany) outside the submitted work. Dr. Bernd Saugel has received institutional restricted research grants and honoraria for giving lectures from CNSystems Medizintechnik GmbH (Graz, Austria) outside the submitted work. Dr. Bernd Saugel is a consultant for and has received institutional restricted research grants from Retia Medical LLC. (Valhalla, NY, USA) outside the submitted work. Dr. Bernd Saugel is a consultant for and has received honoraria for giving lectures from Philips Medizin Systeme Böblingen GmbH (Böblingen, Germany) outside the submitted work. Dr. Bernd Saugel is a consultant for and has received honoraria for giving lectures from GE Healthcare (Chicago, IL, USA) outside the submitted work. Dr. Bernd Saugel was a consultant for and has received institutional restricted research grants from Tensys Medical Inc. (San Diego, CA, USA) outside the submitted work. Dr. Moritz Flick has received honoraria for consulting and giving lectures from CNSystems Medizintechnik (Graz, Austria) outside the submitted work. All other authors report no conflicts of interest.

## Funding

No funding received.

## Acknowledgements

This research was partially supported by the Hellenic Society of Cardiopulmonary Resuscitation, Athens, Greece. The authors would like to thank the medical and nursing stuff of the Department of Anesthesiology, University Hospital of Larisa, for their assistance during the study period. We are also thankful to Z. Hossain, medical software engineer at Microvision Medical (Amsterdam, The Netherlands), who provided expertise that greatly assisted the research.

## Author Contributions

Dr. Athanasios Chalkias designed the study. Dr. Athanasios Chalkias, Dr. Konstantina Kolonia, Dr. Zacharoula Angelopoulou, Dr. Dimitrios Ragias, Dr. Dimitra Papaspyrou, Dr. Nicoletta Ntalarizou, Dr. Aikaterini Bouzia, Dr. Konstantinos Stamoulis, Dr. Aikaterini Kyriakaki, Dr. Eleni Laou, and Dr. Eleni Arnaoutoglou collected the data and performed quality control. Dr. Athanasios Chalkias, Dr. Nikolaos Papagiannakis, and Dr. Eleni Laou analyzed the data. Dr. Athanasios Chalkias, Dr. Bernd Saugel, Dr. Moritz Flick, and Dr. Jesper Eugen-Olsen interpreted the results. Dr. Athanasios Chalkias wrote the first draft of the manuscript. All coauthors provided critical revisions to the manuscript. All authors approved the final version of the manuscript.

## Notes

### Clinical Trial

NCT03851965

### Author Declarations

Ethical approval for this study was provided by the Ethical Committee of the university hospital of Larisa, Greece (IRB no. 60580).

