## SUPPLEMENTARY FILES for "Association of preoperative plasma suPAR levels with intraoperative sublingual microvascular perfusion in patients undergoing major non-cardiac surgery"

**SUPPLEMENTARY FIGURES**

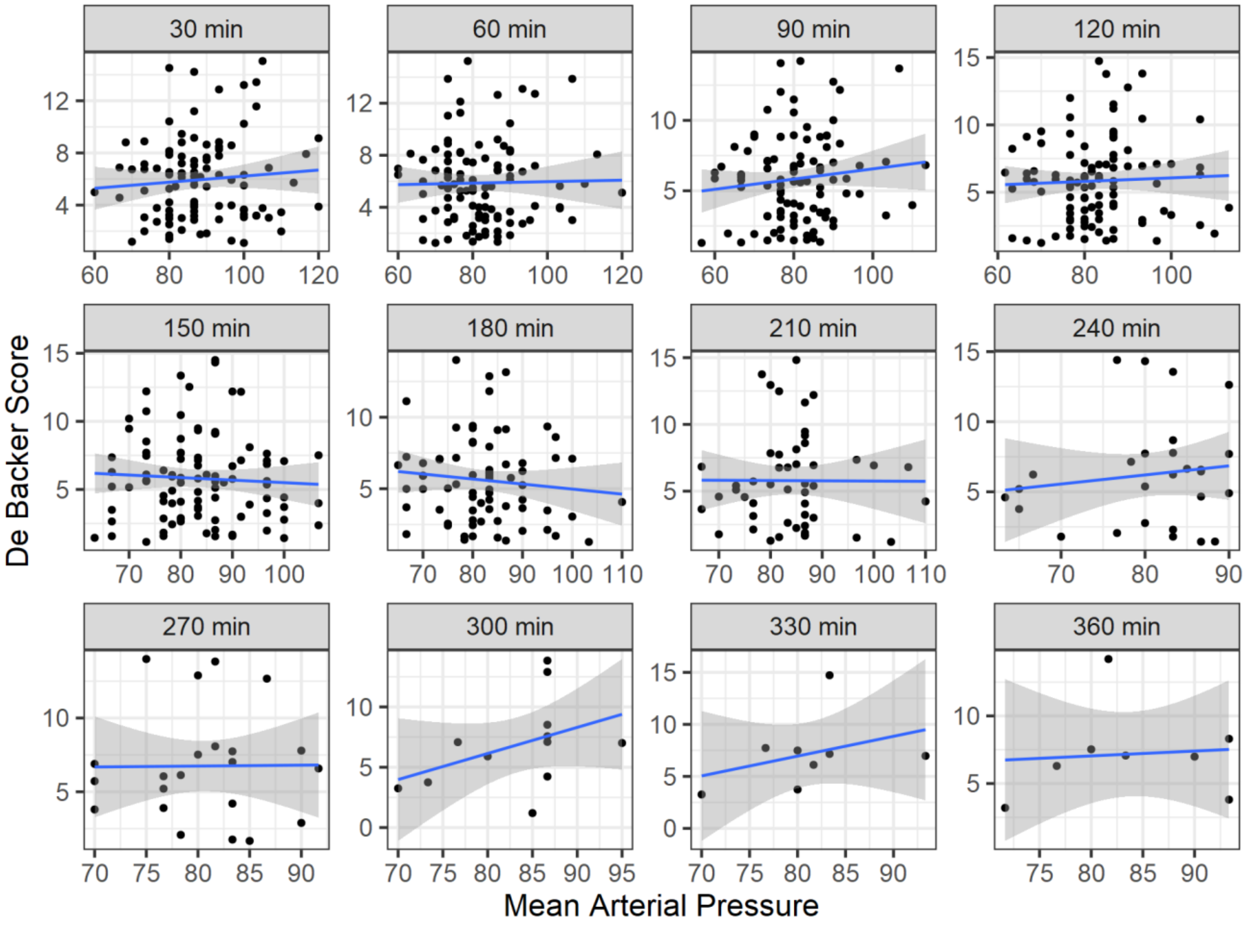


Supplementary Figure 1. Association of mean arterial pressure with De Backer score.

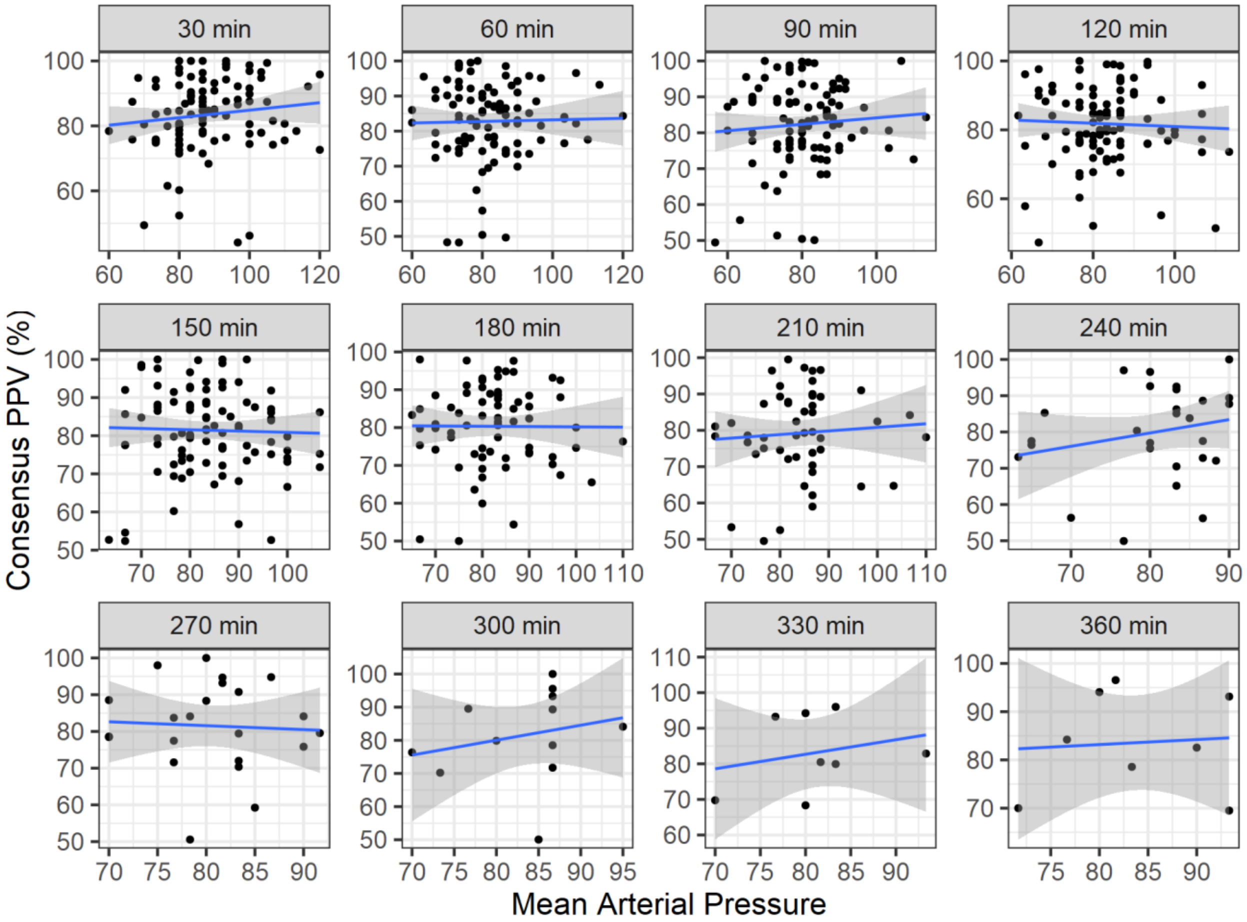


Supplementary Figure 2. Association of mean arterial pressure with Consensus PPV.

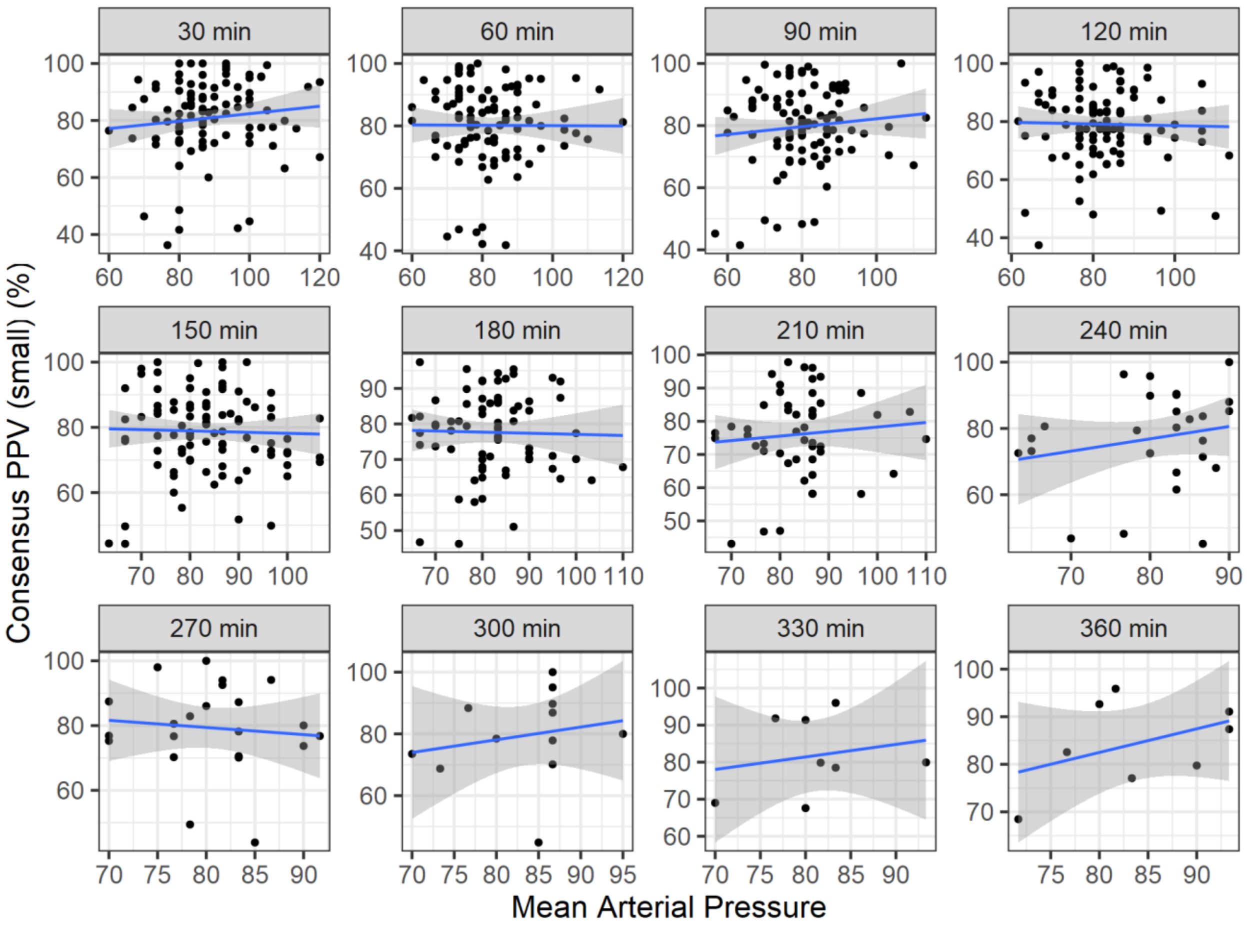


Supplementary Figure 3. Association of mean arterial pressure with Consensus PPV (small).

### SUPPLEMENTARY TABLES

**Supplementary Table 1. Preoperative and postoperative arterial blood gases**

| | Preoperatively | Postoperatively | Adjusted p-values |
|---|---|---|---|
| pH | 7.38 (0.02) | 7.40 (0.03) | <0.001 |
| $PaCO_2$ (mmol/L), mean (SD) | 38.22 (3.3) | 39.88 (2.6) | <0.001 |
| $HCO_3$ (mmol/L), mean (SD) | 23.72 (2.5) | 25.5 (1.8) | <0.001 |
| Lactate (mmol/L), mean (SD) | 1.25 (0.4) | 0.86 (0.2) | <0.001 |
| $SaO_2$ (%), mean (SD) | 97.56 (1.8) | 99.81 (0.6) | <0.001 |
| $SpO_2$ (%), mean (SD) | 97.09 (1.9) | 98.86 (1.2) | <0.001 |

**Supplementary Table 2. Preoperative and postoperative differences in sublingual macrocirculatory flow**

| Variable | Preoperative | Postoperative | p-value |
|---|---|---|---|
| De Baker score | 5.95 (3.21) | 5.89 (3.36) | 0.825 |
| Consensus PPV | 83.49 (11.5) | 81.15 (11.8) | 0.005 |
| Consensus PPV (small) | 80.87 (13.4) | 78.72 (13) | 0.021 |

Data presented as mean (standard deviation)

**Supplementary Table 3. Association between De Backer score, Consensus PPV, and Consensus PPV (small) and systemic hemodynamic variables**

| | Variable | Spearman's rho | Adjusted p-values |
|---|---|---|---|
| De Baker score | Systolic arterial pressure | -0.042 | 0.428 |
| | Diastolic arterial pressure | 0.063 | 0.174 |
| | Mean arterial pressure | 0.026 | 0.584 |
| | Heart rate | -0.020 | 0.678 |
| Consensus PPV (%) | Systolic arterial pressure | 0.000 | 0.998 |
| | Diastolic arterial pressure | 0.032 | 0.547 |
| | Mean arterial pressure | 0,025 | 0.588 |
| | Heart rate | -0.085 | 0.050 |
| Consensus PPV (small) (%) | Systolic arterial pressure | -0.011 | 0.806 |
| | Diastolic arterial pressure | 0.026 | 0.584 |
| | Mean arterial pressure | 0.015 | 0.760 |
| | Heart rate | -0.085 | 0.050 |
